# Automatic phenotyping of electronical health record: PheVis algorithm

**DOI:** 10.1101/2020.06.15.20131458

**Authors:** Thomas Ferté, Sébastien Cossin, Thierry Schaeverbeke, Thomas Barnetche, Vianney Jouhet, Boris P Hejblum

## Abstract

Electronic Health Records (EHRs) often lack reliable annotation of patient medical conditions. Yu *et al*. recently proposed *PheNorm*, an automated unsupervised algorithm to identify patient medical conditions from EHR data. *PheVis* extends *PheNorm* at the visit resolution. *PheVis* combines diagnosis codes together with medical concepts extracted from medical notes, incorporating past history in a machine learning approach to provide an interpretable “white box” predictor of the occurrence probability for a given medical condition at each visit. *PheVis* is applied to two real-world use-cases using the datawarehouse of the University Hospital of Bordeaux: i) rheumatoid arthritis, a chronic condition; ii) tuberculosis, an acute condition (cross-validated AUROC were respectively 0.947 [0.944; 0.948] and 0.987 [0.983; 0.990]). *PheVis* performs well for chronic conditions, though absence of exclusion of past medical history by natural language processing tools limits its performance in French for acute conditions. It achieves significantly better performance than state-of-the-art methods especially for chronic diseases.

## 1. Introduction

As the amount of data collected on a daily basis from hospital health care system keeps increasing,[1] the appeal for leveraging the full potential of these data for research purposes and to investigate clinical questions is also becoming stronger than ever. [2–5] Yet, EHR data are quite different from research oriented data (e.g. cohort or trial data): i) they are less structured, more heterogeneous, ii) they present finer granularity, iii) data collection is done for health care purpose. [1,6–8] Currently, one of the main barriers to use such data for studying disease risk factors is the necessity to first identify patients having diseases of interest, a task that we will denote as phenotyping.

Several approaches have been recently proposed to phenotype patients. [9–13] They often rely on either rule-based algorithms specifically designed with clinicians, or on supervised models trained on annotated patient datasets. Such algorithms are limited because their development is disease specific, must be (re-)started from scratch for every new disease and demand a lot of clinician expertise time. In addition, portability and generalization to new databases (e.g. different hospitals) can often fail, requiring once again the process to be reiterated in the new institution. Hripcsak and Albers defined high-throughput phenotyping as an approach that “should generate thousands of phenotypes with minimal human intervention”. [8] In this perspective, multiple methods have been developed for automatic phenotyping. Agarwal *et al*. proposed *XPRESS* which learns on noisy labels.[10] Halpern *et al* proposed *Anchor* which learns on so-called “anchor patients”, i.e. patients with highly disease-specific features.[11] Wagholikar *et al* developed *Polar*, which learns on so called “polar patients”, i.e extreme patients which are almost certain to either have or not have the disease.[12] Finally Yu *et al*. developed *PheNorm* which learns the phenotype as a continuous score. [13] And they also developed *SAFE* which selects relevant features for phenotyping in an automated manner.[14] All those frameworks are unsupervised, in the sense that they require neither any manual chart review nor any complex rules definitions to classify phenotype, and thus allow automated high-throughput phenotyping.

While those frameworks are appealing, they only consider phenotyping at the patient level and neglect the timing of illness onset and cure. Yet, we need increased resolution for phenotyping, especially for studying acute diseases (that can occur repeatedly) or for answering epidemiological questions (where temporal sequence is important): phenotyping at the visit level would allow to precisely take into account the dynamic evolution of patient’s conditions. Besides, those frameworks were developed using English databases, leveraging advanced NLP tools and relying on rich terminologies not necessarily available in languages other than English. [15,16] Portability to other languages is not straightforward, as they still often lack resources of matching quality.

We propose a new, portable, approach for unsupervised algorithm extending *PheNorm* at the visit level: *PheVis*. This new *PheVis* method cumulate past information to provide an up-to-date estimation of a phenotype probability at any given visit. This accumulation of previous information from EHR can be tuned to match the disease length, making *PheVis* a versatile tool suitable for both chronic and acute conditions. Section 2 presents the *PheVis* analysis and modeling strategy. Section 3 demonstrate *PheVis* performance for rheumatoid arthritis (RA) and tuberculosis (TB), a chronic and an acute condition respectively, using French EHRs from Bordeaux University Hospital. The method is compared to other state-of-the-art methods. Finally Section 4 discusses these findings and the limits of the approach, and offers a conclusion.

## 2. Material and methods

*Phe Vis* combines ICD10 (international classification of diseases 10^th^ revision) billing codes together with medical concepts extracted from clinical notes, incorporating past information through a user-tunable exponential decay. This creates a silver-standard surrogate of the medical condition of interest. Then variable selection (through elastic-net logistic regression) and pseudo-labelling (using random-forest) are performed, leveraging extreme values of this silver-standard. Finally, a logistic regression model is estimated on those noisy labels to provide an interpretable “white box” predictor of the occurrence probability for a given medical condition at each visit. The different steps of *PheVis* are outlined in Figure 1 and are described below.

**Figure 1:**
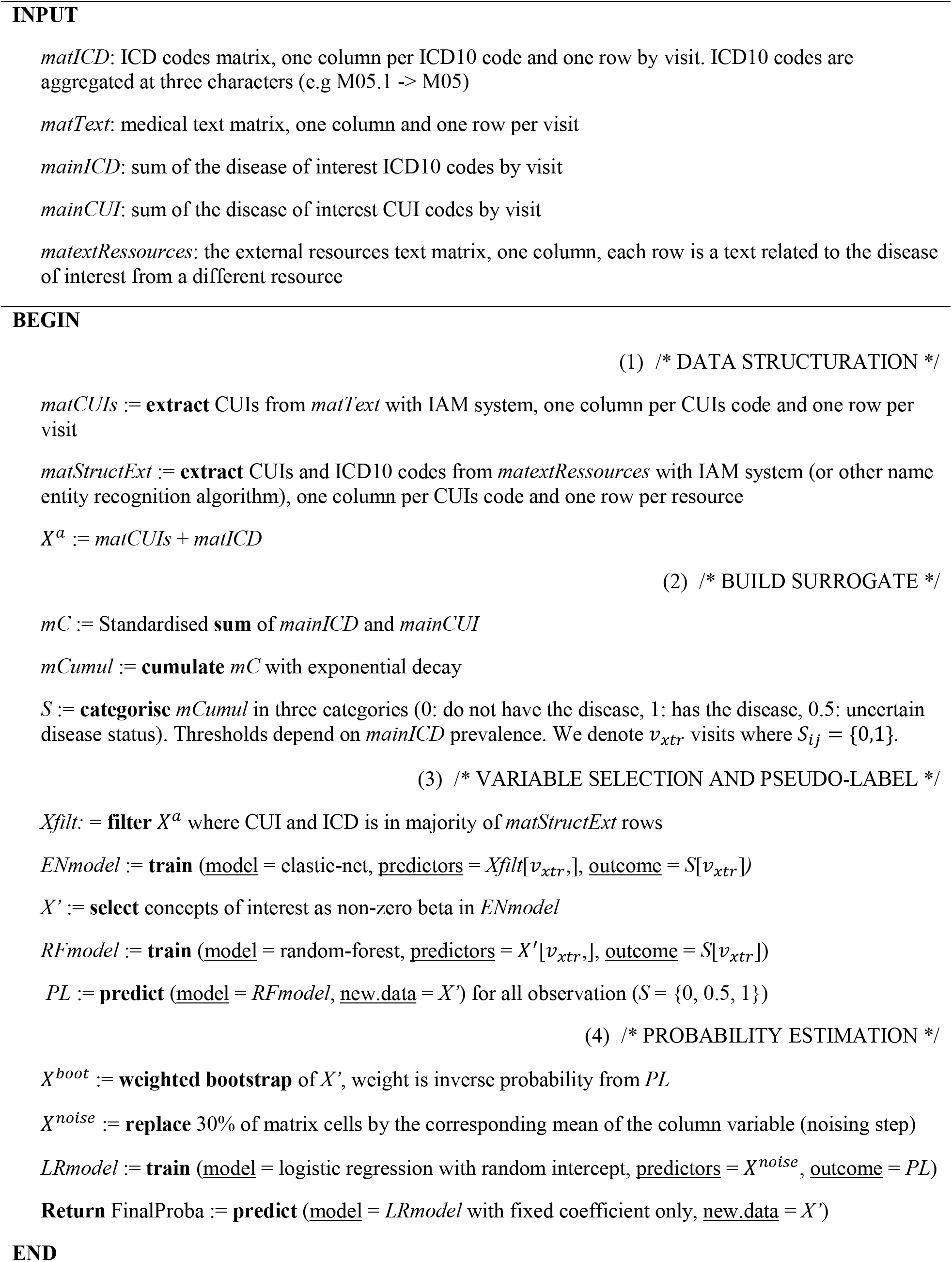
Pseudo-code of PheVis.

### 1. Input data

The input data of the *PheVis* approach are the clinical notes and the ICD10 codes from an EHR datawarehouse. All the notes and ICD10 codes are collapsed by visit, and IAMsystem, a dictionary-based named entity recognition tool, is used to extract relevant UMLS concepts unique identifiers, CUIs (i.e. CUIs associated with disease to be phenotyped – see Section 2.3 for details). [17,18] ICD10 codes are aggregated at the category level (i.e. the first three characters, M05.1 and M05.2 codes are both counted under the same category code M05). This result in a matrix *X* of dimension *φ* × *P*, where *φ* is the total number of visits and *P* the total number of ICD and CUI concepts. We will denote *i* ∊ {1,…, *n*} the patient index and *j* ∊ {*v*_1_, …,*v_i_*} the visit index.

### 2. Build a surrogate of the disease status

As there are no disease labels for the visits (hence requiring a phenotyping algorithm), a supervised model cannot be trained right away. To be able to train our phenotyping algorithm, we first build a surrogate variable expected to be close to the true disease status. This surrogate is based on the main ICD and UMLS codes that represent a disease.

We define *mC_ij_* the standardized sum of main disease concepts as:

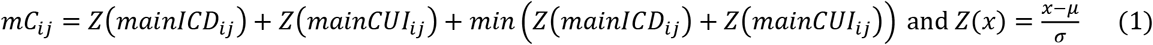

*mainlCD* and *mainCUI* are main concepts related to the disease. For example, for RA we used:

- *mainlCD_ij_* = *M*05*_ij_ + M*06*_ij_* with *M*05*_ij_* the number of times the code *M05 Rheumatoid arthritis with rheumatoid factor* was recorded for patient *i* at visit *j*, and similarly for *M*06*_ij_* and M06 *Other rheumatoid arthritis*
- *mainCUI_ij_* = *C*0003873y with *C0003873 Rheumatoid arthritis*

At this stage, standardization (centering and scaling) is critical because CUIs occurrences often largely outnumber ICD code occurrences. Without such standardization, the weight of ICD codes in the prediction would be negligible.

To phenotype a given visit, it is necessary to take into account information available from previous visits as well. For example, a patient can be diagnosed RA at the age of 50, have a visit at 52 for an infectious event containing no information about RA. RA being a chronic disease, we want to be able to predict RA in both visits. To do so we propose to cumulate past history information with an exponential decay as follow:

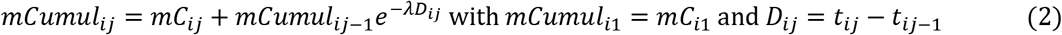

*λ* is a constant parameter tuned by the user that controls the “memory loss” of the algorithm. For easier interpretation one can prefer to set the value of half-life equals to *ln* (2)/*λ*. The natural half-life chosen is the usual duration of the disease (e.g. 180 days for TB and +∞ for RA — being a chronic disease currently without a cure). Setting +∞ for RA is equivalent to simply cumulate the information of all previous visits.

The same exponential decay accumulation is applied to each ICD and UMLS codes. We also define five other features:

- *lastvis_ij_* = *mC_ij_*_−1_
- *last*5*viS_ij_* = 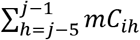
- *lastmonth_ij_ =* 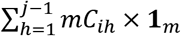 with **1***_m_* = 1 if *D_ij_* − *D_ip_* ≤ 30*days*, 0 otherwise
- *lastyearij =* 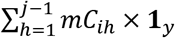 with **1***_y_* = 1 if *D_ij_* − *D_ip_* ≤ 365*days*, 0 otherwise
- *Cum_ij_* = 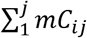

This yields an augmented matrix *X^a^* of *φ* × (2*P* + 5) dimensions: CUIs and ICDs (*P*), their cumulated counts (*P*), and 5 new variables.

## 3. Variable selection and pseudo-labelling

We use the *SAFE* algorithm to select predictive variables of interest and reduce the dimensionality of the optimization problem. First we use IAM system to extract ICD10 and UMLS concepts in external resources: medical text books and Wikipedia disease specific chapter or page.[19–22] A concept and its cumulative count are kept in the model only if it is found in the two resources. Then we categorize *mCumul_ij_* into *S_ij_* = {0,0.5,1}.

To define the two thresholds separating the three categories of *S_ij_*, we used *mainICD_ij_* which takes into account prevalence variability depending on the disease and the cohort. We first count the proportion of visit with at least one occurrence of *mainICD* code (i.e the *mainICD* ≥ 1 prevalence) that we denote *quant_mainICD_* as:

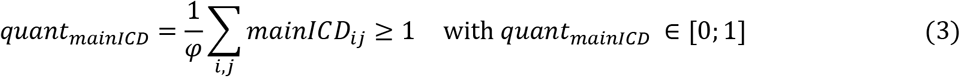

We divide this quantity by a constant *ω* that we set to 2 to define *quant_extreme_* as:

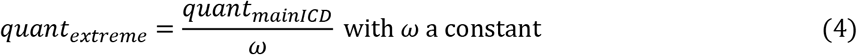

This *quant_extreme_* proportion allows to define three categories: [0; *quant_extreme_*], ]*quant_extreme_*; 1 − *quant_extreme_*[, [1 − *quant_extreme_*; 1] and we define the surrogate *S_ij_* as:

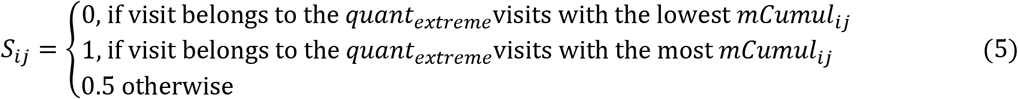

One can note that the higher the *ω* constant (i.e the extreme patients are more extreme), the more confident we are in the specificity of *S_ij_* in {0,1} toward the true phenotype but the smaller training size is for next steps. We found *ω* = 2 to work well in our setting.

Then we train a logistic regression with elastic-net penalization to select a subset *X*′ of relevant predictors from the *X^a^* matrix using only the visits for which *S_ij_* is either 0 or 1. *X*′ is the subset of predictors with non-zero estimated coefficients. Of note, *mainICD* and *mainCUI* are always forced into the set of selected variables in *X*′, while *Cum_ij_* is systematically removed for acute conditions.

We then assign a pseudo-label {0,1} to all visits. This increases the number of visits available to train the final logistic regression, and also adds visits with more uncertain phenotype status which overall results in smoother predicted probabilities and better performance. To perform this pseudo-labelling, we train a random-forest with majority vote for trees aggregation for which *S_ij_* is either 0 or 1. The trained model is then used to predict the pseudo-label *PL_ij_* = {0,1} status for each visit.

## 4. Probability estimation

To estimate the disease occurrence probability, we used a noising-denoising logistic regression with random intercept similarly to *PheNorm*. First, *max*(10^5^, *φ*) visits are randomly sampled with replacement with inverse probability weighting depending on *PL_ij_* in order to balance the training set. This new matrix is denoted *X^b^*. Then we perform a noising-denoising step to force the algorithm to use other variables than the main ICD and UMLS concepts (and thus avoid overfitting with respect to the surrogate construction). Every value of explanatory variables has a probability of *p_bern_* = 0.3 to be replaced by the mean of the explanatory variable as in PheNorm.[13] For instance if M05 ICD10 code mean occurrence is 0.2 then each visit has a probability of 0.3 to have its true M05 value replaced by 0.2. This noisy matrix is denoted *X^n^*:

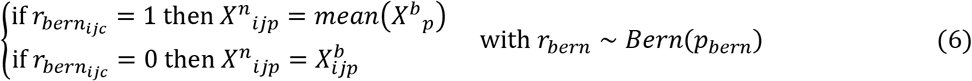

For the denoising step a logistic regression with random intercept is used:

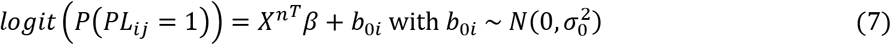

And finally the probability of having the disease is estimated on the noise free matrix as:

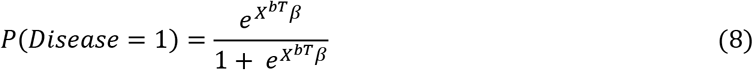

This final probability illustrates the level of confidence of the estimated phenotype based on the used variables.

## 3. Results

### 3.1. Application design

We illustrate the *PheVis* method on RA, a chronic disease which cannot be cured, and active TB, an acute disease which usually last between 6 to 12 months. [19–22] The model performance was evaluated on an imperfect gold standard for both diseases: for RA we used the presence of at least one rheumatoid arthritis form, a form specifically used by rheumatologists at the University Hospital of Bordeaux in usual RA care, for tuberculosis we manually reviewed patients with at least one mention of tuberculosis treatment while other patients were considered not having the disease. Latent tuberculosis was labelled as tuberculosis negative because, even if the bacterium is the same, symptoms, diagnosis and treatment are different. Patients were included if i) they had been hospitalized at the University Hospital of Bordeaux at least once since 2010 and ii) if they had either one primary or secondary ICD code of RA (M05 or M06), or one biology measurement of Anti-Citrullinated Peptide Antibody. The cohort was split into training and test datasets at patient level with a 70% to 30% ratio. The cohort is described in Table 1, highlighting the discrepancy between ICD, CUI and gold-standard justifying the need for automated phenotyping algorithms.

**Table 1.** Description of phenotyping cohort. *University Hospital of Bordeaux*.

|  | Train set |  | Test set |  |
| --- | --- | --- | --- | --- |
|  | Patients | Visits | Patients | Visits |
| n | 9102 | 237875 | 2359 | 62004 |
| Gold standard PR (%) | 27,077 (11.4) | 953 (10.5) | 7,883 (12.7) | 274 (11.6) |
| ICD RA<sup>1</sup> ≥ 1 (%) | 21,448 (9.0) | 3,682 (40.5) | 5823 (9.4) | 901 (38.2) |
| CUI RA ≥ 1 (%) | 32,775 (13.8) | 3,703 (40.7) | 8,632 (13.9) | 952 (40.4) |
| Gold standard TB (%) | 618 (0.3) | 49 (0.5) | 90 (0.1) | 5 (0.2) |
| ICD TB<sup>2</sup> ≥ 1 (%) | 277 (0.1) | 88 (1.0) | 50 (0.1) | 15 (0.6) |
| CUI TB ≥ 1 (%) | 2,393 (1.0) | 647 (7.1) | 439 (0.7) | 147 (6.2) |

1: ICD RA: M05, M06
2: ICD TB: A15, A16, A17, A18, A19

Ten different prediction models were evaluated for each disease: (i) our proposed *PheVis* approach, for which we set 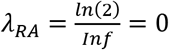 and 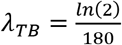 (tuberculosis typically lasting around 6 months), (ii) *XPRESS* for which *Inf* 180 we defined the silver standard as the main ICD code presence, (iii) *ANCHOR* for which we defined the anchor visits as the one with at least one main ICD and main CUI code, (iv) the elastic-net and (v) the random forest *POLAR* algorithms for which we adapted the polar patient definition to our setting (negative polar visits neither main ICD codes and neither main CUI codes, whereas the positive polar visits had both), (vi) *PheNorm*, (vii) a supervised elastic-net with cumulated variables (denoted as *ENETCUM)* and also viii) without cumulated variables *(ENET_NOCUM)*, (ix) a supervised random forest with cumulated variables *(RF_CUM)* and (x) without cumulated variables *(RF_NOCUM)*. Of note, *ENETCUM* and *RF_CUM* can be considered as oracle predictors as they are supervised and learn on both cumulated and non-cumulated variables. We used *SAFE* to select variables used for prediction by each algorithm in order to ease their comparison.

### 3.2. Application results

Figure 2 shows individual *PheVis* and other state-of-the-art methods predictions for four patients. For RA, as the information is cumulated over time in *PheVis*, the model is able to maintain relatively stable predictions over time even if there is not much information about RA in a visit. Without this feature, other approaches display highly variables predictions oscillating over time. For TB, the advantage of cumulating information is confronted to the problem of cumulating too much. This problem is increased in French because the majority of natural language processing tools and terminological systems were developed for the English language. Tools to detect negation and past history are not yet implemented in Bordeaux University Hospital datawarehouse.[23] Prediction of *PheNorm* is really close to 0.5 for all visits. This can be explained by the final step of the *PheNorm* algorithm which is a mixture model on the predicted sum of main ICD and main CUI. As our dataset is largely imbalanced, with many more negative patients than positive ones, both normal distributions of the mixture model are concentrated on the negative class. Because they are close to each other, the probability of belonging to each class (positive or negative phenotype) is really close. However, as shown on figure 3 and 4, those probabilities are not constant and still give good prediction performance according to AUROC and AUPRC, in spite of being poorly interpretable. Other methods do not have this problem, as they mainly learn on binary silver standard.

**Figure 1:**
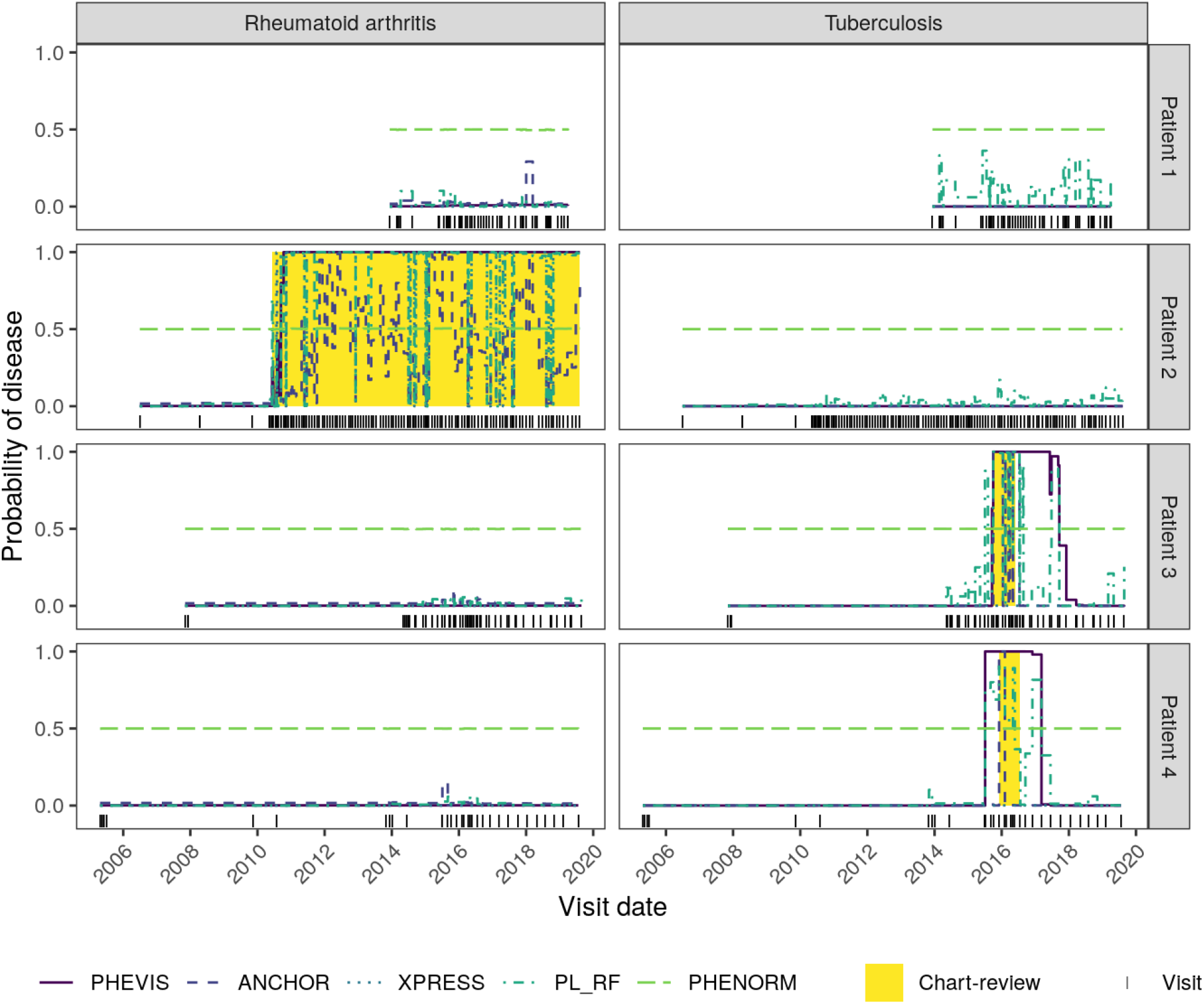
Individual prediction of rheumatoid arthritis (RA) and tuberculosis (TB). Each column corresponds to a disease, each row to a patient. Patient 1 has no disease. Patient 2 has RA which is well estimated by *PheVis*, other algorithms have high variability in their prediction. Patient 3 and 4 have tuberculosis. For both of them, that information is still trailing behind after the patient is cured for *PheVis*, partly because of the lack of advanced natural language processing tool hindering the distinction between past and actual disease history. *PheNorm* predictions are close to 0.5 because the mixture model fails to learn meaningful probabilities in this extremely imbalanced setting. *XPRESS* fails to estimate any probability higher than 0 for TB because of convergence failure and RA probabilities are either almost 0 or 1 providing a binary interpretation of the disease status rather than a continuous one. Both *PL_RF* and *Anchor* are too volatile to provide a trusted interpretation of their output probability.

**Figure 2:**
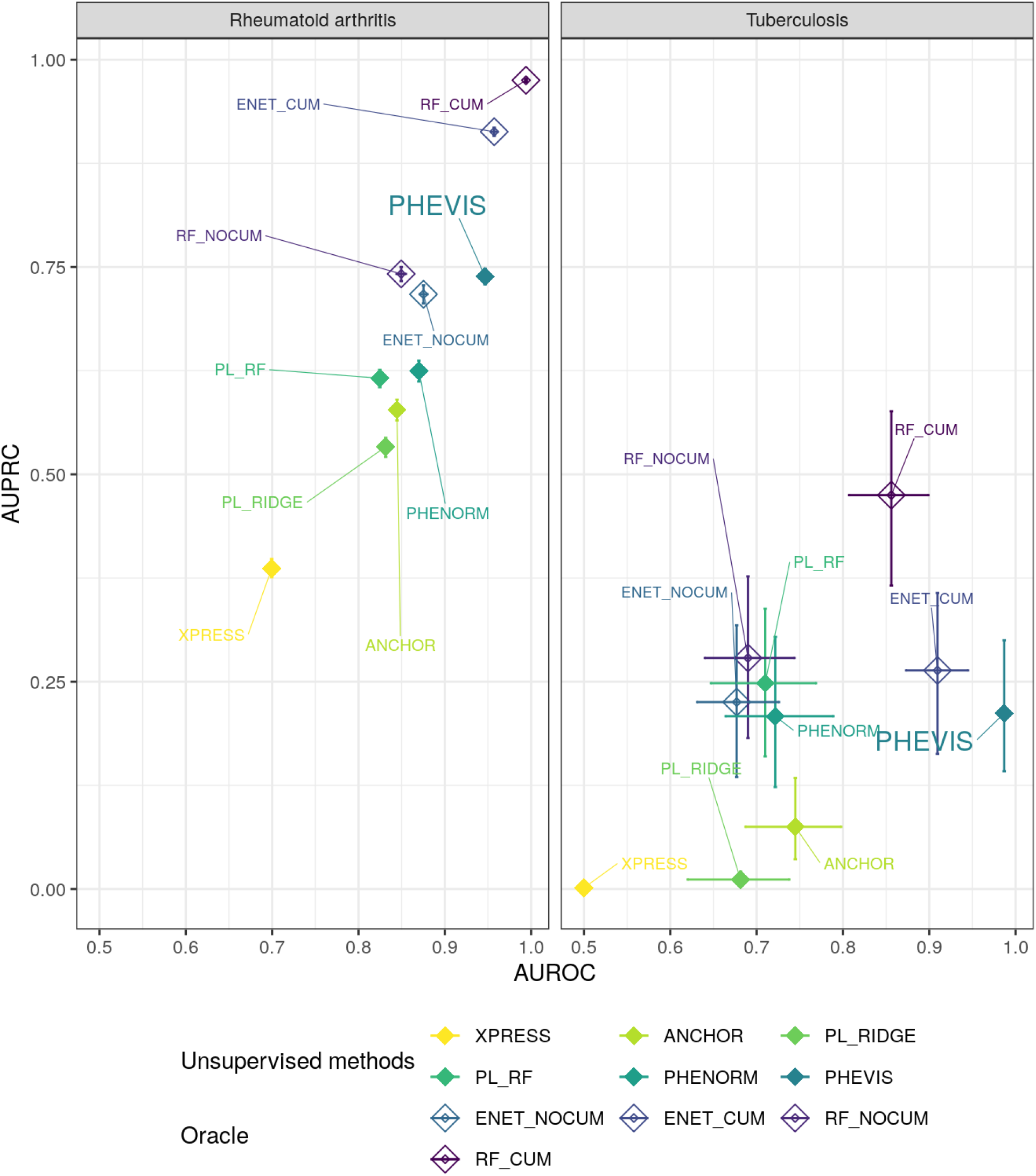
Phenotyping unsupervised and supervised methods comparison. University Hospital of Bordeaux. Confidence Intervals for AUROC and AUPRC are represented by horizontal and vertical segments respectively. PL_RIDGE: *POLAR* method with ridge logistic regression. PL_RF: *POLAR* method with random forest. ENET: Supervised elastic-net logistic regression with (CUM) or without (NOCUM) cumulated variables. RF: Supervised random forest with or without cumulated variables.

Figure 3 shows the performance of *PheVis* and the other methods on the test set for both TB and RA. For RA, *PheVis* brings significantly better performance than any other unsupervised method, both in term of AUROC (RA: 0.947 CI [0.944; 0.948], TB: 0.987 CI [0.983; 0.990]) and AUPRC (RA: 0.739 CI[0.729; 0.748], TB: 0.212 CI[0.142; 0.300]). Table with detailed performances of the algorithms is available in the supplementary material. As expected, for TB the advantage of *PheVis* is less important because the information is cumulated over a shorter time period, still it performs well especially in term of AUROC compared to other unsupervised methods. Supervised methods worked significantly better with cumulated variables supporting the importance of taking into account past history to predict actual phenotype. Among the other unsupervised methods, we can denote that *PheNorm* and the random forest *POLAR* seem to be the best methods, however, as shown on figure 2 *PheNorm* has serious calibration problem on unbalanced datasets. *XPRESS* failed to reach convergence in our setting possibly because it uses lasso penalization instead of elastic-net as in *SAFE* or *PheVis*, or ridge as in *ANCHOR* or *POLAR*. Both for *POLAR* and the supervised “oracle” methods, random forest was able to significantly improve the performance of the model, probably because it is able to learn more complex structure than penalized linear regression.

Table 2 shows point performance for two arbitrary phenotyping decision rules: i) a predicted probability above 0.5 ii) a probability above the threshold maximizing the sum of the precision and the recall. Specificity and negative predictive values are good partly because the diseases are rare at the visit level. Matching the results from figures 3 and 4, the sensitivity/positive predictive values trade-off is better for RA than for TB.

**Table 2.** *PheVis* performance for different thresholds.

| Disease | Threshold | SE | SP | PPV | NPV |
| --- | --- | --- | --- | --- | --- |
| Rheumatoid arthritis | 0.5 | 0.828 | 0.903 | 0.554 | 0.973 |
|  | Optimal P-R*<br>(0.941) | 0.771 | 0.932 | 0.622 | 0.965 |
| Tuberculosis | 0.5 | 0.556 | 0.994 | 0.124 | 0.999 |
| | Optimal P-R*<br>($2.10 \cdot 10^{-5}$) | 0.989 | 0.948 | 0.027 | 1.000 |

\* Threshold maximizing the precision recall sum.

## 4. Conclusions

We developed *PheVis* as an unsupervised automatic phenotyping algorithm at the visit level. Our innovative approach resembles the human medical probabilistic approach of diagnosis as the output is a probability taking into account the uncertainty of the information inside EHR. [24] It is able to achieve good performances for RA, which is promising for other chronic conditions. While *PheVis* represent a significant improvement over the current state-of-the-art thanks to its versatile and tunable information cumulating feature, it also suffers from limitations when it comes to acute conditions such as TB. The algorithm is fully automated, not requiring any (time-consuming and expensive) manual chart review, and can in theory be used for different kind of medical conditions (either acute or chronic).

*PheVis* adds many innovations to the previous *PheNorm* algorithm it builds upon: the needs for standardizing the information from medical notes and ICD codes, the accumulation of past history with exponential decay, the definition of silver standard using ICD codes to take into account prevalence of the disease, and pseudo-labelling to improve performance and increase stability of predicted probabilities. Also we demonstrated the portability (and limitations) of those methods in French and in a different datawarehouse than the one used to develop *PheNorm*, with consistent performances for phenotyping RA compared to Yu et al. [13]

Our application setting is different from the other methods original paper, mainly because there is intra-patient correlation between visits phenotype. This is not accounted for in *Phenorm*, *XPRESS* or *POLAR* where the learning is at patient level, nor in *ANCHOR* because it learns on acute diseases.[10-13] As in *POLAR* or *ANCHOR*, our dataset is largely unbalanced towards the negative class (e.g 1.1% epilepsy prevalence in *POLAR*, 2.0% septic shock prevalence in *ANCHOR)* contrary to *PheNorm* (lowest disease prevalence was RA with 22.5%). This unbalanced setting seems to favors unsupervised learning with silver standard *(PheVis, POLAR, ANCHOR,XPRESS)* in terms of calibration. In terms of performance, *PheVis* performed better for both diseases on both AUROC and AUPRC. Behind, the hierarchy between random forest *POLAR* and *PheNorm* is not clear-cut. As those three methods rely on different approaches, future developments might be able to leverage and combine each of their strengths.

These phenotyping algorithms are highly sensitive to the input features, which emphasizes the needs for finer natural language processing tools able to perform semantic analysis. The use of other features such as biological test results or treatment should also be considered, as they should be highly predictive of the phenotype, but further works is needed to define how they could be integrated into the silver-standard surrogate strategy used in *PheVis*.

Our performance evaluation is made against an imperfect gold standard, mainly due to the lack of large annotated patient reference sets. For TB, the gold standard was manually curated, while for RA, we used a highly specific form but which might lack sensitivity: interestingly, upon manual inspection it appeared that *PheVis* was able to accurately recover RA patients visits of 5 patients who were not treated in the Rheumatology department of the University Hospital of Bordeaux and thus had no record of this specific form, resulting in a failure of the gold standard. Such phenomena might underestimate the algorithm performance.

*PheVis* can provide a probability for a large set of diseases and medical conditions with little effort. The performances might vary depending on the disease of interest, the database quality and the EHR language but were better than state-of-the-art method in our study. The use of those estimated probabilities opens new horizon for the use of EHR for medical and epidemiological research purposes.

## Data Availability

Data were provided by the CHU de Bordeaux.

## Supplementary material

### ICD10 codes

**Table S1:** Main ICD codes of rheumatoid arthritis and tuberculosis used by PheVis.

|  |  |
| --- | --- |
| Tuberculosis | A15, A16, A17, A18, A19 |
| Rheumatoid Arthritis | M05, M06 |

### Algorithm performance

**Table S2:** Phenotyping unsupervised and supervised methods comparison. University Hospital of Bordeaux. PL_RIDGE: *POLAR* method with ridge logistic regression. PL_RF: *POLAR* method with random forest. ENET: Supervised elastic-net logistic regression with (CUM) or without (NOCUM) cumulated variables. RF: Supervised random forest with or without cumulated variables.

| Algorithm | Rheumatoid arthritis |  | Tuberculosis |  |
| --- | --- | --- | --- | --- |
|  | AUPRC | AUROC | AUPRC | AUROC |
| ANCHOR | 0.578 [0.565 ; 0.590] | 0.845 [0.840 ; 0.850] | 0.075 [0.036 ; 0.134] | 0.745 [0.687 ; 0.798] |
| ENET_CUM | 0.913 [0.908 ; 0.918] | 0.957 [0.954 ; 0.961] | 0.264 [0.163 ; 0.357] | 0.909 [0.873 ; 0.945] |
| ENET_NOCUM | 0.717 [0.706 ; 0.728] | 0.875 [0.871 ; 0.880] | 0.225 [0.135 ; 0.318] | 0.677 [0.631 ; 0.726] |
| PHENORM | 0.625 [0.612 ; 0.637] | 0.870 [0.865 ; 0.875] | 0.208 [0.123 ; 0.304] | 0.722 [0.664 ; 0.789] |
| PHEVIS | 0.739 [0.729 ; 0.748] | 0.947 [0.944 ; 0.948] | 0.212 [0.142 ; 0.300] | 0.987 [0.983 ; 0.990] |
| PL_RF | 0.616 [0.605 ; 0.626] | 0.825 [0.819 ; 0.831] | 0.248 [0.160 ; 0.338] | 0.710 [0.647 ; 0.769] |
| PL_RIDGE | 0.533 [0.521 ; 0.544] | 0.832 [0.826 ; 0.837] | 0.011 [0.006 ; 0.021] | 0.681 [0.620 ; 0.738] |
| RF_CUM | 0.975 [0.973 ; 0.977] | 0.994 [0.993 ; 0.995] | 0.475 [0.366 ; 0.576] | 0.856 [0.807 ; 0.899] |
| RF_NOCUM | 0.742 [0.733 ; 0.750] | 0.850 [0.844 ; 0.855] | 0.279 [0.182 ; 0.377] | 0.690 [0.640 ; 0.744] |
| XPRESS | 0.386 [0.376 ; 0.398] | 0.699 [0.695 ; 0.705] | 0.001 [0.001 ; 0.002] | 0.500 [0.500 ; 0.500] |

